# Machine learning models for early prognosis prediction in cardiogenic shock

**DOI:** 10.1101/2025.09.18.25336054

**Authors:** Nicolás Nieto, Federico Raimondo, Simon B. Eickhoff, Janine Pöss, Steffen Desch, Hans-Josef Feistritzer, Artur Lichtenberg, Maryna Masyuk, Malte Kelm, Holger Thiele, Kaustubh R. Patil, Christian Jung

## Abstract

Cardiogenic shock (CS) is a severe and frequent complication of acute myocardial infarction (AMI), necessitating rapid and accurate prognosis assessment to guide treatment and intensive care unit (ICU) resource allocation. We developed two machine learning models to predict 30-day outcomes following CS in AMI: an Admission model (using only data available at admission, like demography, comorbidities) and a Full model (incorporating additional laboratory values obtained within 24 hours). The models were trained on the CULPRIT-SHOCK dataset and externally validated using the eICU database. The Admission model achieved an out-of-sample AUC of 0.71 (95% CI: 0.6–0.83) in the development cohort and 0.68 in the validation cohort, while the Full model attained significantly higher performance, with AUCs of 0.80 (95% CI: 0.69–0.9) and 0.78, respectively. The Full model’s superior performance underscores the prognostic value of early laboratory trends, suggesting that dynamic data integration improves risk stratification. Both models outperformed existing risk scores across multiple metrics, provided well-calibrated probabilistic predictions, and demonstrated robustness to missing data. Additionally, they offered patient-level explainability, enhancing clinical interpretability. While promising, the models’ generalizability may be influenced by differences between the CULPRIT-SHOCK and eICU cohorts (e.g., demographics, CS severity thresholds); further validation in larger, prospective cohorts is warranted.

## 1. Introduction

Cardiogenic shock (CS) is a syndrome caused by a primary cardiovascular disorder in which inadequate cardiac output can result in multi-organ dysfunction and death.^1^ The most common cause of CS is acute myocardial infarction (AMI). The annual incidence of CS is more than 30,000 patients in the United States and 45,000 in Europe, with a mortality rate close to 50%.^2, 3^ Cardiogenic shock is a complex and heterogeneous condition, encompassing a wide spectrum of clinical presentations and severities, which can vary across different patient phenotypes.^4^ To address this variability, the Society for Cardiovascular Angiography and Interventions (SCAI) has proposed a standardized classification system that categorizes CS into stages (A through E) based on clinical and hemodynamic criteria.^5^ This framework aims to provide a unified language for describing CS severity, facilitating consistent communication among clinicians and researchers across diverse settings. However, while this system provides a valuable descriptive tool, it does not offer quantitative risk stratification or mortality prediction.

Fast and accurate estimation of a patient’s prognosis can improve treatment and allow for better allocation of the scarce resources in the Intensive Care Unit (ICU). To this end, several risk scores have been proposed, such as the Intra-Aortic Balloon Pump SHOCK II score (IABP-SHOCK II),^6^ Simplified Acute Physiology Score (SAPS II),^7^ Shock trial registry score,^8^ CardShock score,^9^ BOS,MA_2_ score,^10^ and Cystatin C, Lactate, Interleukin 6, NTproBNP (CLIP) score,^11^ among others. However, these scores present two main limitations, impairing their utility in real-world applications. First, they use laboratory test values, which are not readily available at admission, limiting their application in real scenarios, as risk assessment is needed as soon as possible, ideally within minutes after patient admission. Moreover, some of the scores rely on information that might not be broadly available at every ICU, like Cystatin C or Interleukin 6. Second, these scores cannot handle missing values, which are of common occurrence in a clinical setting as data is obtained under critical circumstances, and other problems, such as laboratory equipment not functioning and misplaced records, may occur. Therefore, robust models that can make accurate predictions, even with missing information, are desirable.

Machine learning (ML) models have been used to integrate diverse patient information to obtain an accurate and fast diagnosis in different clinical scenarios in general,^12, 13, 14^ and in CS in particular.^15, 16, 17, 18^ However, such models have not yet been explored for the prognosis of CS. An appropriate ML model can assist clinical decision-making by providing several advantages; (1) prediction at the time of admission can help make decisions regarding the trajectory of the care given to a patient, (2) providing an uncertainty estimate along with a prediction, (3) handling of missing information, and (4) patient-level explainability of how the model arrives at a prediction. ML models are trained using patient information (called features), like demographics (age, sex), electrocardiogram results (heart rate, sinus rhythm, atrial fibrillation), and laboratory test results (creatinine, lactate, troponin), among others. A trained model can predict a newly admitted patient’s characteristics, expressed as a target, for example, the presence of a disease or the patient’s prognosis. In the training phase, ML models use already acquired and labeled data to automatically adjust their internal parameters to learn the relationships between the features and the target. When a new patient arrives, the trained ML model can make a prediction based on the same features for this new patient.

In a clinical setting, it is important to obtain uncertainty estimates together with a prediction. Indeed, some classification models can provide such an output. However, it must be ensured that this output is calibrated, meaning that the model’s uncertainty estimate refers to the actual likelihood of an event happening. For instance, the model is correctly calibrated if it predicts a 70% probability of a patient presenting a disease, and, on average, 70% of the patients predicted with that probability presented the disease. A well-calibrated model is of vital importance where the decision-making relies on the model’s output, as miscalibrated models can lead to suboptimal or harmful treatment decisions.^19^

In this work, we propose an ML approach that satisfies the aforementioned criteria. The proposed models were named MACRO (MAchine learning models for early prognosis prediction in CaRdiogenic shOck). Specifically, we aimed to develop two models, one that only uses features available at admission and a second model that uses features typically available within the first 24 hours after admission. These models were extensively tested for their accuracy, usefulness in uncertainty estimation, and robustness to missing values. Finally, we aim to develop an easy-to-use web service that allows clinicians to use the proposed models for new patients in an intuitive way.

## 2. Methods

### 2.1. Data description

Our study used the CULPRIT-SHOCK dataset,^20^ a high-quality dataset of a prospective multi-center trial, to train the proposed models. This dataset was collected between April 2013 and September 2015 in over one hundred centers across Europe. The dataset was originally designed to compare a culprit lesion-only percutaneous coronary intervention (PCI) with an immediate multivessel PCI regarding the primary composite endpoint of 30-day mortality or renal failure requiring renal replacement therapy. One of the noteworthy characteristics of this dataset is that the data was validated, which means that an expert supervised each loaded value. This provides a high signal-to-noise ratio, a low number of wrongly inputted data, and high data completeness. The dataset included 686 patients (162 females, age mean 68 - standard deviation 11 years) (Supplementary Figure 1). Thirty-day mortality was used as the target, and the dataset presents 361 “Alive” and 325 “Expired” patients.

Missing values were not imputed, as imputed data might not match the underlying data distribution and can compromise the interpretability of classifier models trained using poorly imputed data.^21^ Additionally, one of the main assumptions of most common imputation methods, like the Multiple Imputation by Chained Equations (MICE),^22^ is the Missing at Random (MAR). This assumption requires that missing values appear randomly in the data and are not correlated with the target of interest. Imputing the data when it is not MAR could result in biased estimations.^23^ In our dataset, this assumption is not fulfilled, as the “Expired” patients presented a higher percentage of missing values in the features, compared with the “Alive” patients (Supplementary Table 1 and 2, and Supplementary Figure 2).

The eICU dataset^24, 25, 26^ was used as a validation cohort. This dataset presents 139,367 unique patients who were admitted to 335 different ICUs at 208 hospitals located in the United States between 2014 and 2015. In this dataset, ICU admissions for all causes were collected; therefore, CS patients only represent a small proportion of the whole dataset. Of the total number of patients, 10,337 were admitted to a Cardiac ICU. From this cohort, 245 were diagnosed with CS (ICD: 785.51) and were selected as the validation cohort. According to the dataset information, the cause of CS was myocardial infarction for 30 patients, therefore, broader CS causes can be expected in this dataset. The definition and criteria for defining CS vary across studies,^27^ thus other CS cohorts may be extracted from the eICU dataset.

In contrast with the CULPRIT-SHOCK dataset, the eICU dataset was not validated, meaning that the data registered had no human supervision. This is especially important for those features derived from the bedside vital signs monitors, like heart rate, systolic, and diastolic pressure.

For both datasets, written informed consent was obtained at data acquisition time, and only de-identified data was used.

### 2.2. Admission features

Twenty-five features were selected by two experts due to their availability at the admission time and potentially containing relevant information about patients’ prognosis, such as age, height, weight, heart rate, and blood pressure. These features were selected as they are expected to be available in any ICU setting.

Following the experts’ recommendation, we combined three variables, “Previous myocardial infarction”, “Previous PCI”, and “Previous CABG surgery”, into one unified “Known coronary artery disease”. This variable was set as “True” when at least one of the constituent variables was “True” and was set to “False” otherwise. From all the admission features, six features with a variance less than 0.1 were removed, as the features were almost constant and provided no information. Removing low variance features helps reduce the model’s complexity, mitigating overfitting and improving model explainability.^28^ This resulted in 19 features for the “Admission” model with only a few missing values (each feature had, on average, 1.5% of missing values), as these features are easily accessible (see Supplementary Table 1 for details).

The selected admission features were also extracted from the eICU dataset. Values before admission were used for all variables except for heart rate, systolic, and diastolic blood pressure, for which the information from the first 30 minutes after admission was used. The median of the available measurements, after removing biologically implausible values outside the range from 20 to 200, was computed as the admission value for these features. Notably, only 20% of the patients had blood pressure information, and 80% had Heart Rate (HR) information. The extracted features in the eICU dataset presented a higher number of missing values compared to the features extracted from the CULPRIT-SHOCK dataset (Supplementary Table 2).

### 2.3. 24 hours after admission features

The same two experts also selected another 20 features that are typically measured in the first 24 hours after admission and may be broadly available in different ICUs, such as creatinine, lactate, and hematocrit, among others (Supplementary Table 3). In the CULPRIT-SHOCK dataset, laboratory test results have a time-stamp, and only the features available within the first 24 hours were used, discarding any information that was collected after that point. Additionally, since these results were measured in different units at each site, a unit harmonization was performed. For each feature, the values were multiplied by a conversion factor depending on the needed unit conversion. For example, white cell counts were harmonized into values between 0.1 and 60 10^9^/L.

Five features with less than 0.1 variance were eliminated, resulting in a total of 15 24-hour features. These features had a higher number of missing values compared to admission features (each feature had, on average, 22% of missing values). The details of the 34 features used for the Full model, together with the number of missing values, unit, and variance, are shown in Supplementary Table 3.

Importantly, 90 patients who expired within the first 24 hours were excluded from the cohort from which the Full model was trained. This choice was made to match the training cohort with the patients to whom the model will be applied, as the Full model is designed to be applied only to patients who have survived 24 hours after admission. Additionally, the inclusion of those patients could lead to a biased model as those patients do not have 24 hour lab values. Consequently, a model would learn that if a feature like “lactate 24hs” is not available, then the patient has expired. If the “lactate 24hs” would not be measured for any other reason for a new patient who survived the first 24 hours, the model will predict “Expired” as a prognosis with high likelihood, erroneously influenced by the absence of this feature.

The selected 24-hour features were also extracted from the eICU dataset. The features were harmonized to match the units for the CULPRIT-SHOCK dataset, and the values between admission and the first 24 hours were used. For each feature, if several measurements were acquired in the first 24 hours, the mean of all the values was computed. The extracted features presented a much higher number of missing values, compared with the CULPRITSHOCK extracted features (Supplementary Table 4).

### 2.4. Risk scores

The proposed models were compared with four conventional risk scores calculated in the CULPRIT-SHOCK dataset: CardShock, CLIP, SAPS II, and IABP-SHOCK II, and with the BOS,MA_2_ score in the eICU dataset. As all the risk scores needed laboratory values to be calculated, we excluded the same 90 patients who expired in the first 24 hours for a fair comparison with the Full model.

The SAPS II and IABP-SHOCK II scores are already available in the CULPRIT-SHOCK database for 541 and 409 patients, respectively. These scores were not recalculated. It was possible to calculate the CLIP score for 374 patients, following the procedure in.^11^ It was possible to calculate the CardShock score for 141 patients of the CULPRIT-SHOCK dataset (patients who presented information in all the features needed for the score). It was possible to calculate the BOS,MA_2_ score in 144 of the 245 patients in the eICU cohort, who presented information in all the features needed to calculate the score. It is important to note that the features and weights used for the CLIP and the BOS,MA_2_ scores were obtained in a data-driven fashion using the CULPRIT-SHOCK and eICU datasets used here, respectively.

It was not possible to calculate the BOS,MA_2_, or other general risk scores, like the quick Sepsis Related Organ Failure Assessment (qSOFA) score,^29^ in the CULPRIT-SHOCK dataset, as some of the features that compose the scores are not available.

The SAPS II score uses 15 features, while CardShock uses 7, IABPSHOCK II and BOS,MA_2_ use 6, and the CLIP score uses 4 features. The details of the features that compose each of the compared risk scores are presented in Supplementary Tables 5, 6, 7, 8, and 9.

### 2.5. Machine learning approach

The eXtreme Gradient Boosting (XGBoost)^30^ was used as it provides a fast and efficient way to deal with different data types, as clinical data present a mix of integer (age, heart rate, height, weight), boolean (sex, diabetes mellitus, atrial fibrillation) and float values in different ranges (lactate, creatine, white cell counts). This method has been successfully used to predict CS risk in ST-elevation myocardial infarction (STEMI) patients^15^ or early prediction of CS clinical data.^16^ Another key characteristic of XGBoost is the ability to deal with missing values,^30^ and several tools have been proposed to assess model explainability and feature importance.^31, 32, 33, 34^ The proposed models were trained to predict the 30-day mortality as the target.

The “Admission” model was trained using the 19 features available at admission time, while the “Full” model also used 15 features (34 in total) measured in the first 24 hours after admission. All patients were used for the Admission model, while the 90 patients who died in the first 24 hours were removed from the cohort used by the Full model.

To evaluate the performance of the models, stratified cross-validation (CV) was used. This approach involves dividing the dataset into multiple subsets, or “folds”, and then training and testing the model multiple times, each time using a different fold as the test set and the remaining folds as the training set. In our experiments, the dataset was divided 100 times using a stratified 10 times repeated 10-fold CV scheme. For each split, we followed the same workflow to train and evaluate both models (Figure 1). The stratification in the CV avoids biases by ensuring the same proportion of Alive and Expired patients in each fold. Hyperparameter tuning was performed using OPTUNA,^35^ using slightly stricter hyperparameter ranges as proposed in^36^ (Supplementary Table 10). We used 100 OPTUNA trials, using the default sampler (TPESampler) and a median pruner to eliminate non-promising trials.^36^

**Figure 1:**
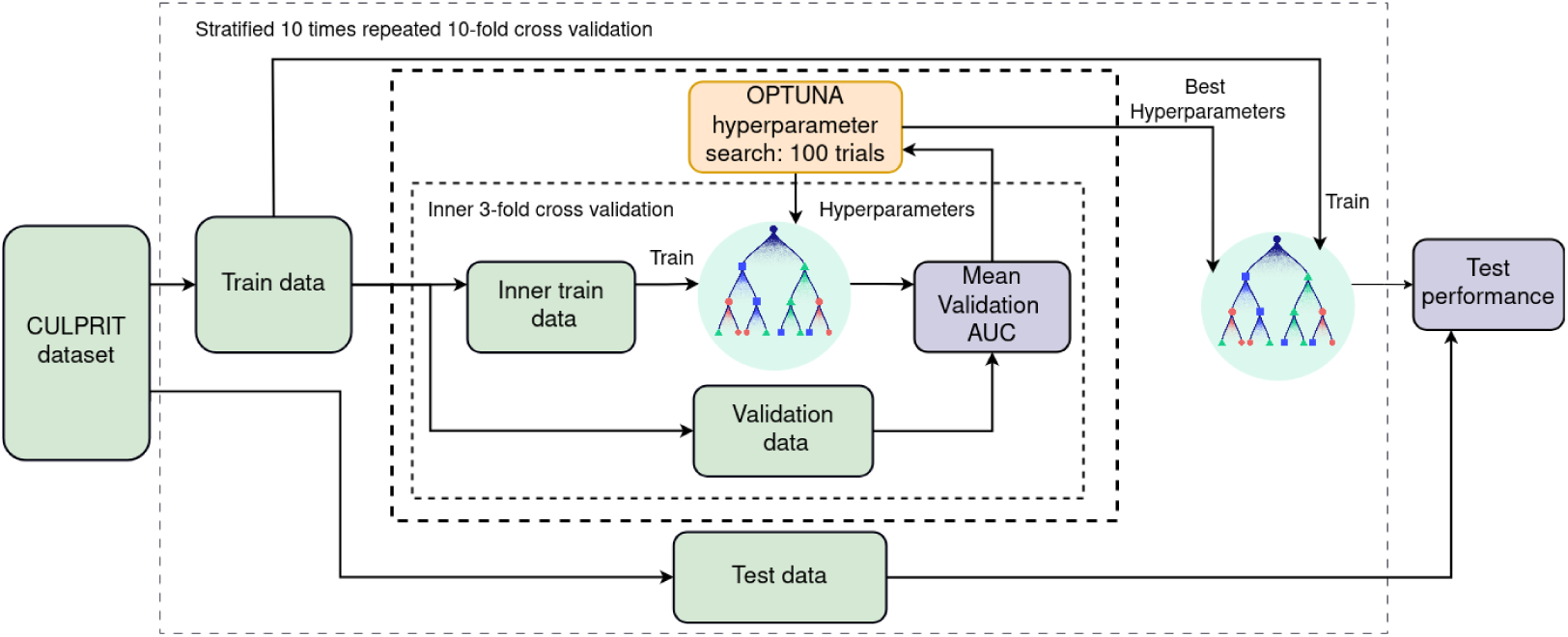
Overall Workflow for each split. The following workflow was used for both the Admission and the Full model. The entire dataset was split 100 times into Train and Test using a stratified 10-fold x 10 repetitions cross-validation. For each train partition, 100 combinations of hyperparameters were sampled using OPTUNA. For each combination, the best number of trees was tuned using a stratified 3-fold inner cross-validation. Once the hyperparameter combination that maximizes the performance over the validation split was obtained, a model was trained using the selected parameters over the entire training data. That model was then used to generate a prediction over the test data, which will later be compared with the true labels to create the performance metrics.

Additionally, to further prevent overfitting typically observed in tree-based ensemble models, we used early stopping with 100 rounds to obtain the number of estimators. To that end, the train data was split into inner train and validation data, and the training procedure stopped when no improvement in a given criterion was achieved on the validation set. The area under the receiver operating characteristic curve (AUC) was used as the optimization criterion. This procedure was repeated using an inner 3-fold stratified CV, and the mean AUC was used for OPTUNA as an optimization goal. The hyperparameter combination that achieved the best mean validation AUC was then used to train a model on the whole training data, which was then applied to the test data (Figure 1). The model’s probability output (continuous value between 0 and 1) was transformed to a binary label (Alive/Expired), using the standard threshold of 0.5.

### 2.6. Score models

To compare our proposed models with conventional risk scores, we built one model per score using logistic regression (LG).^37^ For these models, the 90 patients who expired in the first 24 hours were removed, as all the scores require laboratory data. This cohort matches the cohort used for the Full model.

Each score model takes as input one feature: the SAPS II score, IABPSHOCK II score, CardShock, or the CLIP score, and predicts 30-day mortality. The same stratified 10 times repeated 10-fold CV scheme used to evaluate the proposed models was also used for the score models. In this case, those patients who had missing information (where the scores were not calculated or could not be calculated) were removed from the train and test folds. The BOS,MA_2_ score has a risk probability associated with each of the possible score values, which was used as model output (Supplementary Table 9).

### 2.7. Metrics

The balanced accuracy (bACC), sensitivity, specificity, harmonic mean of the precision and recall (F1), and AUC were computed to extensively compare the performance of the proposed models and the risk-score-based models. For each metric, the 95% Confidence Interval (CI) with equal areas around the median was calculated. The “Expired” class was used as the positive class. All metric formulas (if applicable) and implementation are shown in Supplementary Table 11.

### 2.8. Random permutation test

The random permutation test is a statistical technique commonly used in ML to assess the significance of a model’s performance by comparing its classification performance to a distribution of possible outcomes obtained through random shuffling of the data. This test checks if the model captured the real feature-target relationship. In our experiments, all the labels were randomly permuted before splitting the data in train and test, following the recommendations in.^38^ The models’ training and hyperparameter search were performed in the same fashion as in the main experiments, but now using the shuffled data. Before each CV, the data was shuffled. By repeating this 100 times, we obtained a total of 1000 performance values using randomized labels.

### 2.9. Missing values analysis

To systematically study the impact of the missing values, several experiments were performed to understand the models’ behavior with different amounts of missing information. The ability to make a prediction even when some of the information is missing is one of the key characteristics of our model, as this is expected in real clinical settings.

For this purpose, the Admission model was trained with all available data as previously described, but some of the test sample features were gradually removed while age and sex were assumed to be always available.

The same procedure was followed for the Full model, i.e., trained the model with all available data, but the test sample features were gradually removed. In this case, all admission features were assumed available to the model. The feature removal was done in three ways: i) Random scenario: randomly removing different numbers of features for each patient; ii) Worst case scenario: Removing the most important features first. For instance, creatine was removed first, then lactate 8hs, lactate 16hs, and so on; iii) Best case scenario: The features were removed in the inverse order of their importance. The importance of the features was obtained using Shapley Values,^31, 32^ as described in Section 2.11.

### 2.10. Missing values baseline

Aiming to quantify the extent the models use the missing value for predictions, an experiment was conducted using only the missing value patterns. Here, all available feature values were replaced with 0, and all missing values were replaced with 1, creating a dataset that contains the missing value patterns and effectively uses only the presence or absence of data as input. This approach aims to isolate the contribution of missing value patterns to the models’ predictions, as the models could identify systematic missing value patterns and use this bias to achieve an accurate prediction.

If the models do not rely on missing value information, their performance should be at the chance level (AUC = 0.5). This experiment was applied to both the Admission and Full models, following the same training and evaluation methodology as in the main analysis. The results establish a baseline performance of the influence of missing values on the models’ predictive performance, which can serve as a comparison to determine to which extent the missing values could bias the models’ predictions.

### 2.11. Feature explanation

Not all the features provided to the models carry the same information about a given patient’s prognosis. Some features are more important than others, having different impacts on the model’s performance. Moreover, features for each patient can impact the model’s decision differently; thus, individual-level explainability is of crucial importance in clinical applications.

In our study, Shapley Values^31, 32^ were used to explore the features that influence the model’s output at the patient level. This method computes the feature importance for each sample by comparing the model’s prediction with or without using a particular feature. This contribution, called SHAP values, is positive or negative depending on whether the feature value pushes the model’s decision toward Expired or Alive, respectively. In addition to providing a patient-level explanation for a decision, the averaged absolute SHAP value across individuals provides a measure of a feature’s importance.

To evaluate redundant features, the hierarchical clustering implemented in the shap library was used (https://shap.readthedocs.io/en/latest/. Briefly, the method clusters the features by training XGBoost models to predict the outcome for each pair of input features. The distance between clusters represents the similarity between features, where a distance of 0 represents identical features and a distance of 1 represents independent features.

## 3. Results

### 3.1. Models performance

The mean and 95% CI of the 10 repetitions of 10-fold CV obtained on the CULPRIT-SHOCK, for each metric and model, are presented in Table 1. Even though the IABP-SHOCK II score showed good performance in other cohorts,^6^ the score showed no clear relationship with 30-day mortality in the CULPRIT-SHOCK dataset (Supplementary Figure 3). The model that used this score was not able to extract useful information and was biased toward predicting the samples as Alive, resulting in chance-level balanced accuracy, specificity 1, and sensitivity 0 (Table 1). We, therefore, excluded this score from further analysis.

**Table 1:** Averaged results obtained on the cohort original CULPRIT-SHOCK dataset from the 10 folds x 10 repetitions. The mean and 95% confidence intervals (CI) are presented for each metric and each model. Abbreviation: MV=Missing Values

| Model | bACC (%) | AUC | F1 | Specificity | Sensitivity |
| --- | --- | --- | --- | --- | --- |
| Admission model | 64.6<br>[53.1/76.1] | 0.715<br>[0.598/0.832] | 0.618<br>[0.483/0.753] | 0.687<br>[0.53/0.844] | 0.605<br>[0.432/0.778] |
| Full model | 70.7<br>[59.2/82.2] | 0.799<br>[0.69/0.908] | 0.633<br>[0.472/0.794] | 0.817<br>[0.704/0.93] | 0.597<br>[0.395/0.799] |
| SAPS II | 59.7<br>[49.0/70.4] | 0.677<br>[0.536/0.818] | 0.423<br>[0.219/0.627] | 0.858<br>[0.733/0.983] | 0.336<br>[0.13/0.542] |
| CLIP | 66.3<br>[55.2/77.4] | 0.794<br>[0.653/0.935] | 0.534<br>[0.334/0.734] | 0.898<br>[0.791/1.005] | 0.429<br>[0.213/0.645] |
| CardShock | 56.7<br>[37.3/76.1] | 0.694<br>[0.418/0.97] | 0.341 [-<br>0.115/0.797] | 0.807<br>[0.454/1.16] | 0.328 [-<br>0.196/0.852] |
| IABP-SHOCK II | 50 [50/50] | 0.479<br>[0.277/0.681] | 0 [0/0] | 1 [1/1] | 0 [0/0] |
| Admission model MV Baseline | 50.6<br>[46.6/54.6] | 0.508<br>[0.443/0.573] | 0.064 [-<br>0.075/0.203] | 0.977<br>[0.917/1.037] | 0.036 [-<br>0.045/0.117] |
| Full model MV Baseline | 52.7 [44.4/61] | 0.541<br>[0.416/0.666] | 0.251<br>[0.025/0.477] | 0.875<br>[0.732/1.018] | 0.18 [-<br>0.007/0.367] |

The Admission model, the only model capable of generating a prediction at admission time without using any laboratory values, correctly predicted 64.6% of the patient’s prognosis (Table 1). Importantly, on average, the Admission model predicted at “Risk” for 73% of the patients who expired within the first 24 hours, which is the main goal of this model.

The proposed Full model outperformed all the models in the bACC (70.7%, compared to 66.3% for the CLIP score and 59.7% for SAPS II) and obtained an AUC similar to the CLIP score model (0.799) (Figure 2a). Importantly, the Full model showed a good balance between sensitivity and specificity, largely outperforming the risk-score-based models in the F1 score.

**Figure 2:**
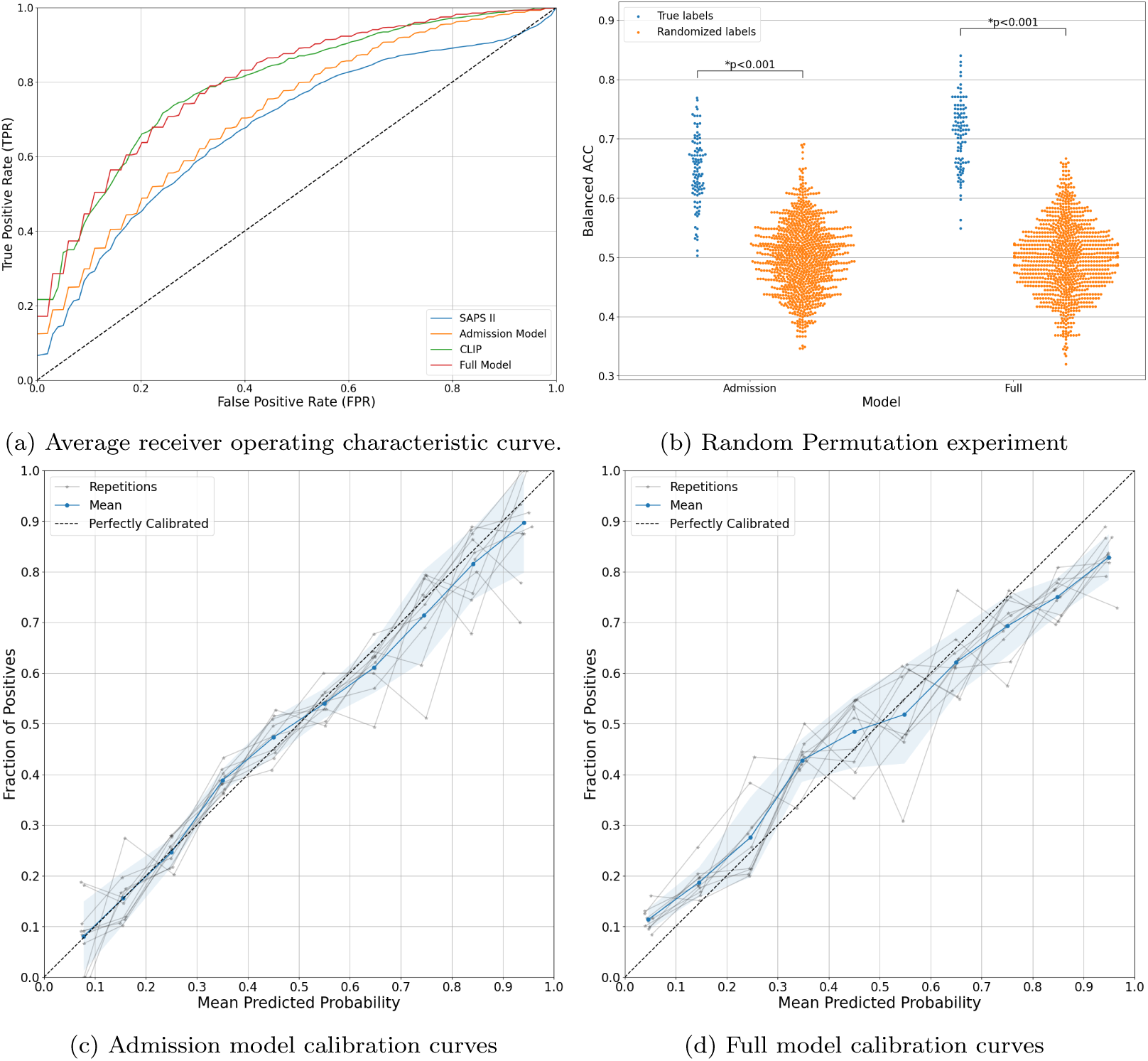
Performance measurements. **a**) Average receiver operating characteristic curve. The True Positive Rate and False Positive Rate were computed and averaged for each fold and repetition. This was repeated for each model. The Admission model performs comparably to the SAPS II model, but it was able to deliver the prediction without requiring any laboratory data. The Full model obtained a similar performance to the CLIP model. **b**) Random Permutation experiment. The balanced accuracy (bACC) for the True and Randomized labels is depicted. For the Randomized labels, 10 different randomizations were performed for each of the 10 folds (no repetitions used). Each point represents the bACC obtained for each randomization in each fold. The blue distributions are obtained in the 10-fold 10 repetitions using the True labels. **c**-**d**) Calibration plots. For each model, the relationship between increasing the model output (probability) and the fraction of positive cases is depicted. For creating each curve, we used the test predictions obtained in the 10 folds of each repetition and compared them with the true labels. Both models presented a good calibration curve, allowing the user to interpret the model output as probabilities. The model shows a good calibration, which can then be approximately considered as the probability of one patient to “Expire”. This is an important characteristic of the machine learning model, mainly in medical domains. Shade areas account for one standard deviation.

The regularization applied to the Full model, even using a restrictive hyperparameter range, was not able to completely avoid overfitting, as the model achieved almost a perfect training performance (Supplementary Table 12).

The permutation test showed that both proposed models’ bACC performance was significantly better when trained using actual labels than when using randomized labels (Mann-Whitney-Wilcoxon test, p < 0.001) (Figure 2b).

The proposed models were well calibrated (Figure 2c). The models presented slightly biased predictions for probabilities close to 0 or 1. For probabilities close to 0, the actual proportion of positive cases was higher than predicted, indicating an underestimation. Conversely, for probabilities close to 1, the actual proportion of positive cases was lower than predicted, indicating an overestimation.

On the validation cohort, extracted from the eICU dataset, both proposed models obtained a similar classification performance, as the one obtained in the discovery dataset (Table 2). For the Admission model, the AUC dropped from 0.712 to 0.68, while for the Full model, the AUC dropped from 0.799 to 0.777. This small drop in performance supports the robust cross-validation scheme used. The BOS,MA_2_ score obtained a better AUC of 0.82 compared with the proposed models and similar to the one reported in the original paper. This is expected, as this score was developed in a data-driven fashion using the eICU dataset. Notably, the Admission model obtained a better F1 score (0.52), compared with the Full model (0.49) and the BOS,MA_2_ (0.32).

**Table 2:** Results obtained on the eICU validation dataset.

| Model | bACC (%) | AUC | F1 | Specificity | Sensitivity |
| --- | --- | --- | --- | --- | --- |
| Admission model | 62.24 | 0.683 | 0.556 | 0.593 | 0.593 |
| Full model | 70.7 | 0.777 | 0.493 | 0.886 | 0.385 |
| BOS,MA<sub>2</sub> | 58.4 | 0.82 | 0.324 | 0.96 | 0.203 |

### 3.2. Missing values analysis

Regarding the Admission model, in the worst-case scenario a steep performance drop was observed when removing dyslipidemia, the most relevant feature after age (Figure 3a). The performance continued dropping and reached chance-level performance when the 9 most relevant features were missing. On the other hand, the model was able to maintain a high performance when 8 features were missing in the best-case scenario. In the random scenario, the performance showed a smoother decline and reached chance-level performance when 9 features were removed.

**Figure 3:**
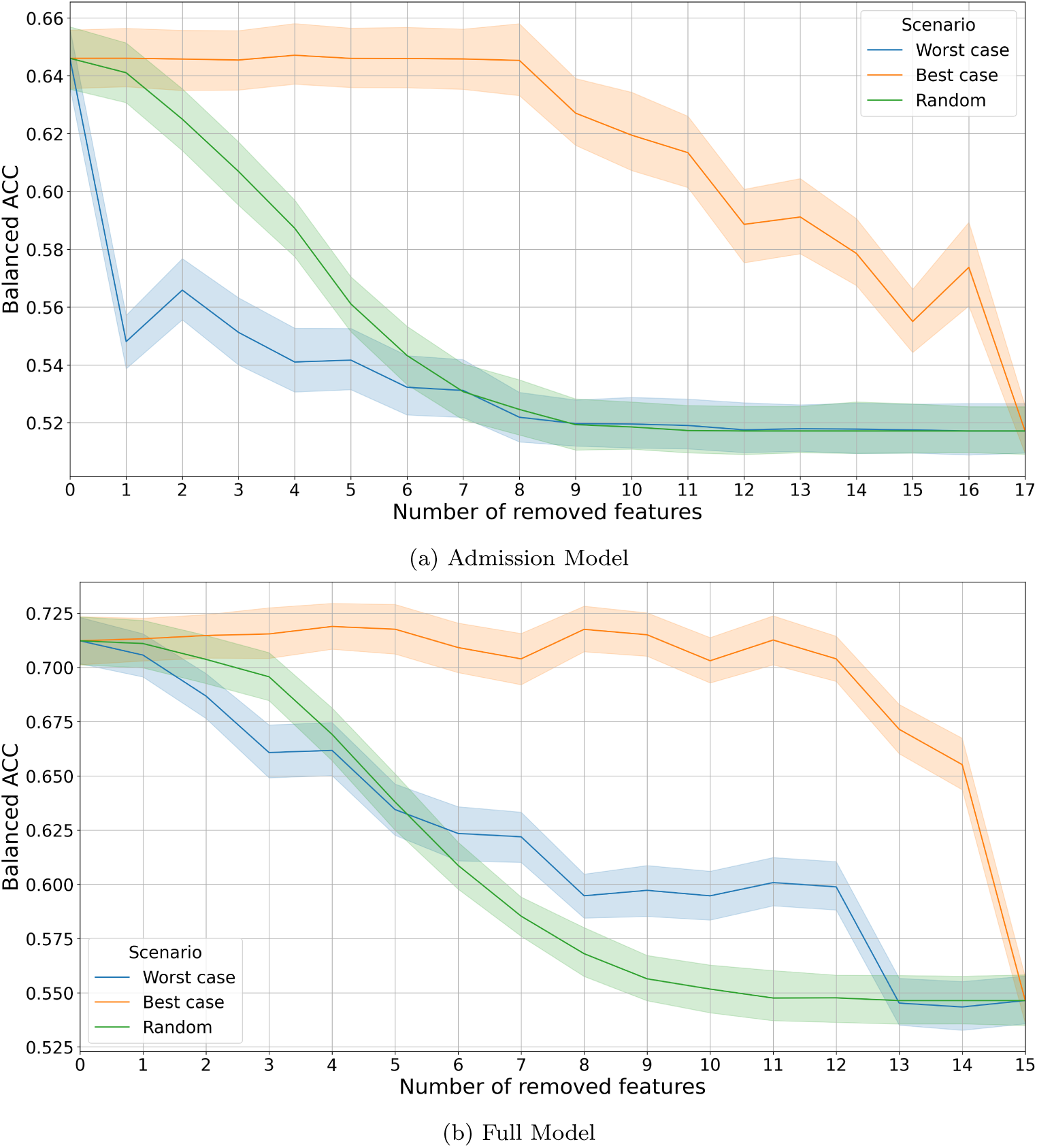
Impact of missing values on the developed models. Features values were replaced with missing values in three different scenarios. In the worst-case scenario, the features were removed from the model following the importance order obtained in the Feature Importance analysis. In the best-case scenario, the features were removed in the inverse of the worst-case scenario. An intermediate scenario, where the features were removed randomly, is also depicted. **a**) Admission model analysis. The 17 admission features besides sex and age, were removed following the different scenarios. For the best-case scenario, the model is able to maintain its performance until 8 features are missing. Compared with the Full model, in the random scenario, the drop in performance is faster. **b**) Full model analysis. The 15 features obtained in the first 24 hours were removed following the described scenarios. In this case, the model always has the 20 admission features available for generating the output. In the best-case scenario, the model can maintain its performance even when 10 features are missing.

In the case of the Full model, a similar behavior to the above-described was observed (Figure 3b). However, this model never reached the chance level, as all the admission features were still available. When the top 4 most informative features were presented (creatine, lactate 16hs, NTproB), the model obtained a similar performance as when all the features were presented (Figure 3b - Best-case scenario with 12 removed features). For the random scenario, the performance after removing 6 features was lower than the Worst-case scenario. This could be due to features that presented higher importance, also encoding similar information. When randomly removing features, different information available to the model was removed, which could lead to stronger detrimental performance.

### 3.3. Missing values baseline

Finally, the Admission and Full models, which were trained using real data, achieved significantly higher AUC scores compared to the models trained using the missing values baseline approach, where missing values were replaced with 1 and real data with 0 (Table 1). The baseline models using missing values yielded AUC scores close to 0.5, indicating classification performance near chance level. The substantial difference in AUC between the real models and the missing values baseline models confirms that classification performance relies on the actual data rather than the distribution of missing values.

### 3.4. Feature importance

For the Admission model, the most important features, calculated as average absolute SHAP values across patients, were age, dyslipidemia, heart rate, and diastolic blood pressure, while the lowest 8 features had a low impact on the model’s output (Figure 4a).

**Figure 4:**
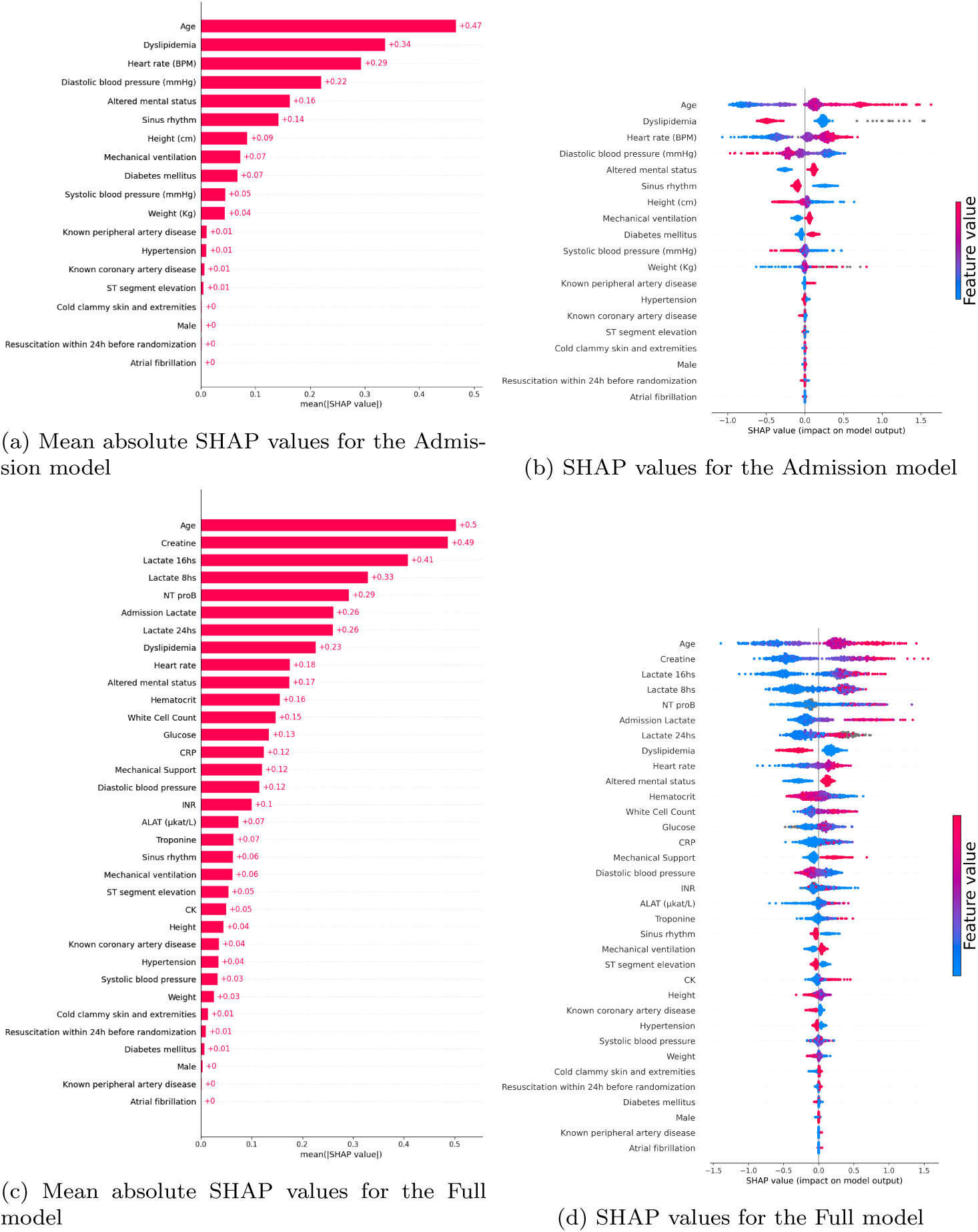
Shapley values for the admission features. To endorse the interpretability of our models, we perform the Shapley values analysis to assess the relative feature importance. **a**-**c**) Mean absolute of the SHAP values for the Admission (a) and 24 hours (c) features. **b**-**d**) Impact of the feature values on the SHAP value for the admission (b) and 24 hours (c) features. For each feature, each point represents a patient in the dataset. When the model output is positive, the model predicts the patient’s prognosis as Expired. The red color is related to high values in the corresponding features, while blue represents lower values. Missing values are depicted in gray dots. Taking Age as an example, when the value of the feature increases (older patients), the feature pushes the model to predict an Expired patient.

For the Full model, the lactate values measured at different time points were the most important features, along with creatine, age, and NT proB (Figure 4c). This result corroborates the CLIP score, which, in a data-driven approach, arrived at similar features using the same dataset. The laboratory values obtained in the 24 hours provide valuable information that the Full model used. Even though age remains a relevant variable, it had a greater influence on the models’ output compared to other laboratory values.

The low impact of some features can be related to the redundancy in the information provided by different features. In essence, the presence of multiple features with overlapping could make the SHAP values of one of them lower. For the Admission model, dyslipidemia is highly correlated with hypertension, but the latter is almost not used by the model. This is also the case for dyslipidemia, diabetes mellitus, and known peripheral artery disease (Supplementary Figure 4). Additionally, other features formed close clusters: diastolic and systolic blood pressure, atrial fibrillation and sinus rhythm, and sex, height, and weight. For the Full model, an expected cluster of lactate values was found. Also, the laboratory values ALAT, INR, and NTproB presented shared information (Supplementary Figure 5). Having redundant information helps maintain the model’s performance even when some features are missing, making the model more robust in the absence of specific variables.

### 3.5. Patient-level explainability

An important aspect of employing ML models in clinical domains is patient-wise explainability, providing a better understanding of how the model arrives at a particular decision. Our model can indeed provide the impact of each feature on each decision.

We assessed the patient-level explainability using the SHAP values “waterfall” plot depicting how the value of each feature contributes to a specific decision for a given patient. For a randomly picked Alive patient, for whom the Admission model correctly classified as Alive, the most important feature that influenced the model to predict the patient as Alive is the age of 41. The rest of the features had a small or no impact on the model’s decision (Figure 5a).

**Figure 5:**
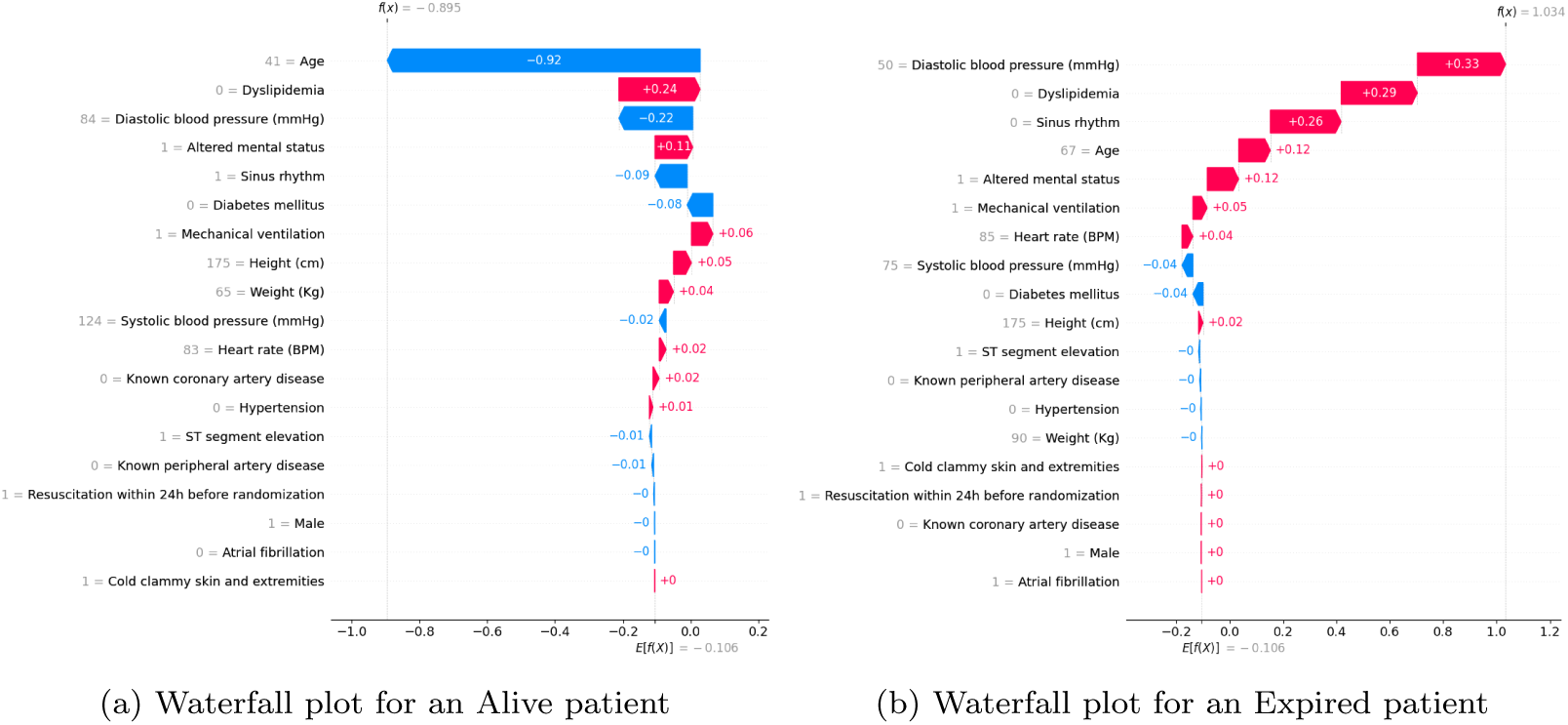
Patient-level explainability. The top f(x) value represents the final model output. Output greater than 0 indicates that the model output was “Expired”, as that class was used as a positive class. On the Y axis, each feature with its corresponding value for a patient is presented. On the X axis, the contribution of the feature value to the model output is presented. Negative values, presented in blue, push the model decision to predict “Alive” patients, while the positive values, presented in red, push the model to predict “Expired” patients. **a**) The waterfall plot from one Alive is depicted. In this case, the Admission model correctly classified the patient as Alive. The most important feature for the output is the Age of 41. The rest of the features have a small or no impact on the model’s decision. **b**) Waterfall plot from an Expired patient whom the Admission model was successfully able to classify. In this case, age does not play an important role in the model’s decision. Heart rate, dyslipidemia, and diastolic pressure are the main factors that push the model’s decision to predict “Expired”. In this case, the height pushes the model decision in the “Alive” direction.

On the other hand, for an Expired patient whom the Admission model successfully classified, age did not play a determinant role in the model’s decision, but rather the Diastolic blood pressure of 50, the absence of dyslipidemia, and absence of Sinus rhythm pushed the model’s decision (Figure 5b).

These two cases exemplify how a clinician can transparently analyze the model’s decision and take this information into account for making a more informed decision.

### 3.6. Web Service

We provide a Web Service demo that can be accessed at: (can’t be provided by double blind). This demo can be publicly accessed and aims to show how clinicians can use the proposed models in a fully functional version. Simulated patients can be loaded in the demo and predictions can be made using both proposed models. Additionally, a patient-wise plot can be obtained for each patient, aiding the interpretation of the output of the model. The demo is designed to showcase the tool’s capabilities while considering legal and privacy concerns associated with decision-making tools in clinical scenarios.

In the EU Artificial Intelligence Act, medical decision-making might be classified as a high-risk application. Therefore, the demo is limited to loading “fake” patient data and does not include the functionality to input values manually.

The fully functional version will allow to input data through manual entry, entailing the completion of the feature values, or via uploading a single.csv file. Most of the features in the Admission model are binary (yes or no), significantly speeding up data entry. Only six numerical variables—age, height, weight, heart rate, systolic and diastolic blood pressure—are used. This minimizes the time required for data entry, facilitating rapid risk stratification even in urgent clinical scenarios. For the Full model, as the 19 admissions features will already be loaded, only the top 4 most informative features (creatine, lactate 16hs, NTproB, and lactate 8hs) need to be input into the model to obtain a similar performance as when all the features are presented. The Web Service automatically validates all features, issuing warnings if a feature is not in its biologically plausible range, mitigating human errors during data input.

## 4. Discussion

ML has gained importance in the field of medicine in the last few years, though studies implementing ML in critically ill patients remain limited. Recently, ML models and other innovative phenotyping technologies have been successfully applied to different cohorts of CS patients,^15, 16, 4^ in addition to several risk scores.^6, 7, 11, 10, 8, 9^ We developed two models, called MACRO models, the Admission model using only admission data, and the Full model using additional laboratory values available within 24 hours. We demonstrated that the proposed ML models can predict a new patient’s prognosis after CS following AMI using a dataset from a large multicentre trial (CULPRIT-SHOCK) and were validated using an external dataset (eICU). Compared to established prognostic scores in CS, the MACRO models are characterized by superior predictive performance. Furthermore, both models fulfill several desired criteria important for clinical application, as they can handle missing information, provide (un)certainty estimates, and provide insights with patient-level explainability. In summary, the proposed MACRO ML models in CS represent strong tools for outcome prediction that allow decision support concerning the potential prognosis of a patient, as early as admission.

The Admission model is the only model able to provide predictions at the patients’ admission, gaining crucial time in the CS treatment. This model uses 19 features broadly available in any ICU at the admission time, like age, heart rate, and comorbidities. This model obtained a cross-validation AUC of 0.71 in the CULPRIT-SHOCK (developing cohort) and demonstrated a good generalization performance, obtaining an AUC of 0.68 on the eICU dataset (external validation cohort). A key strength of this model is that it was able to correctly classify 73% of the “Risk” patients that expired in the first 24 hours. These particular patients are the main goal for the Admission model, as after the first 24hs, the Full model can be applied. Rapidly and accurately labeling a patient at risk can result in better surveillance (e.g. extensive laboratory assessment in shorter time windows), optimizing diagnostics, and evaluating treatment strategies. This model can be potentially valuable to better inform patients and relatives.

Laboratory values are typically used to identify CS and are commonly acquired in a wide range of ICUs, such as lactate, creatinine, and NT-proB. However, some laboratory tests, like Interleukin-6 or Cystatin C, are not broadly available. Conventional risk scores use this laboratory test, which can hinder its use in broad scenarios. To overcome this limitation, the proposed Full model relies on broadly available laboratory tests, making this model suitable for a broad range of hospitals, as no expensive or specialized equipment is needed to obtain the used features. This model outperformed all other models compared here and was able to maintain its performance even with a large number of missing values. The model achieved a high classification performance (AUC of 0.799 and bACC of 71%) when the 5 most important features, besides the admission features, were available. Using the external validation cohort extracted from the eICU dataset, we demonstrated that the Full model obtained a similar performance (AUC: 0.777) compared to the one achieved on the CULPRIT-SHOCK dataset.

Both models showed good calibration and delivered predictions that could be correlated with the frequency of an event occurring, which facilitates and improves the output’s interpretation. However, clinicians and decisionmakers should interpret predictions and associated confidence values with caution, especially when the probabilities are close to 0 or 1.

The range of features included in our models allows for flexibility and robustness in generating accurate predictions, even in cases when some features are missing. This trade-off between the number of features and the model’s ability to handle missing values is crucial. A model with too few features might fail to manage missing data effectively, whereas a model requiring too many features could be impractical for real-time clinical applications. In that regard, the model’s performance did not drop if only the 9 most informative features were available. These characteristics make the Admission model an appealing option, representing the first step to ML-assisted medicine in several ICUs where laboratory tests might be difficult to obtain or have a high cost. From a practical perspective, this model can be applied to generate a broad overview of the patients until more information from laboratory tests becomes available, and the Full model can be applied for a more accurate prognosis.

We would like to note that there is some degree of tolerance for missing values in the admission features. However, we emphasize the importance of the availability of information regarding clinical history and demographics. Importantly, we demonstrated that the good performance obtained by both proposed models was not driven by the missing values presented in the data, as the baseline performance obtained using only missing value information was close to chance.

The patient-level predictions of the models were able to be explained using the shap library. This is an important characteristic of the proposed models, providing not only calibrated and accurate predictions, but also the importance of each feature in the model’s output.

The fully functional Web Service will facilitate prospective users in loading their data and generating predictions utilizing any of the developed models. The automatic checks and easy data entry will mitigate the burden of the number of features used by the proposed models. Furthermore, if the most important variables are loaded in the model, the user can discard the other variables and obtain the same classification performance, speeding up the process. The integration of the patient-level explanation plots provides a fast and broad characterization of each patient.

Regarding the limitations of the present work, several can be acknowledged. First, even when one of the main strengths of our study is the use of a high-quality dataset derived from a large prospective multicenter study, other larger cohorts could be used to train ML models. The decision to use the CULPRIT-SHOCK dataset as a development cohort was based on its high signal-to-noise ratio and relatively low number of missing values, compared with other available datasets. Another limitation is that our model has been mainly tested on CS patients after AMI and validated on a relatively small cohort with broader CS etiology. Further validation needs to be addressed to systematically validate the performance in other larger shock populations. Additionally, not all possible variables known as potential outcome predictors in CS, such as central venous oxygen saturation, have been included in the risk factor analyses. Furthermore, even though developed and tested in a patient cohort from an originally prospective study, our models have not been validated prospectively in CS patients, which remains challenging in the acute setting of critically ill patients. This leads to the general ethical limitation in relying on ML-derived prognoses in medical bedside decision-making, possibly running the risk of retraining treatment for several patient groups. Nevertheless, we suggest here a quick and easy-to-use risk stratification tool, which can be combined with further clinical parameters.

In future works, we aim to generate a more compact model that can deal with missing values without significantly decreasing performance. Furthermore, ML models that take other real-world scenarios into account are desirable. For instance, features often come in “batches” after different medical procedures and laboratory tests. If it is not possible to perform an ECG on the patient, the derived features (sinus rhythm, atrial fibrillation, and ST-segmentation elevation) will not be available altogether.

In conclusion, we developed and validated two ML models that can be broadly applied in a wide range of ICUs and are available via our Web Service. These models provide calibrated uncertainty estimates, can deal with missing values, and generate patient-level explanations. This study may be an important contribution toward ML-assisted decision-making in a clinical setting.

## 5. Data availability

For the CULPRIT-SHOCK, no personally identifiable information was used and individual participant data are not available for sharing. For the eICU dataset, data is publicly available at https://physionet.org/content/ eicu-crd/2.0/ after registration.

## 6. Code availability

All codes were written in Python. Data processing and handling were made using the Python libraries Pandas^39^ (version 1.4.4) and Numpy^40^ (version 1.23.3). We used the implementation of XGBoost available at https://github.com/dmlc/xgboost (version 2.0.2). For all the score models, we used the “LogisticRegression” class implemented in sklearn,^41^ version 1.1.2. The scores calculation was made using the libraries sklearn, imblearn (version 0.10.1^42^), and scipy (version 1.9.1^43^). Shap analysis was performed using SHAP 0.45.1,^32, 31^ and plots were generated using Matplotlib^44^ 3.6.0 and Seaborn^45^ 0.12.0.

The Python environment and all the codes used to perform the experiments and generate the plots in this paper are publicly available at: https: //github.com/double-blind-submission-AIM/MACRO_EXPERIMENTS.

Furthermore, to provide final-user models, one Admission and one Full model were trained using the best hyperparameter range (Range 2) and using 100 OPTUNA trials to obtain the best set of hyperparameters. The final-user Admission model was trained on the entire dataset, while the final-user Full model was trained on the dataset excluding the patients that expired in the first 24 hours. Both models are publicly available in the same repository.

## Data Availability

For the CULPRIT-SHOCK, no personally identifiable information was used and individual participant data are not available for sharing.
For the eICU dataset, data is publicly available at https://physionet.org/content/eicu-crd/2.0/ after registration.

## Acknowledgments

This work was supported by the MODS project funded from the program “Profilbildung 2020” (grant no. PROFILNRW-2020-107-A), an initiative of the Ministry of Culture and Science of the State of Northrhine Westphalia and by the Helmholtz Portfolio Theme Supercomputing and Modeling for the Human Brain. The CULPRIT-SHOCK study was supported by a grant (FP7/2007-2013) from the European Union 7th Framework Program and by the German Heart Research Foundation and the German Cardiac Society.

## 7. Author Contributions

NN: conceptualization, data curation, formal analysis, investigation, methodology, software, validation, visualization, writing – original draft, and writing – review and editing

FR: conceptualization, formal analysis, investigation, methodology, resources, software, supervision, validation, visualization, writing – original draft, and writing – review and editing

SE: conceptualization, formal analysis, funding acquisition, investigation, methodology, project administration, resources, supervision, writing – review and editing

JP: conceptualization, funding acquisition, methodology, resources, supervision, review and editing

SD: conceptualization, funding acquisition, methodology, resources, supervision, review and editing

MM: conceptualization, funding acquisition, methodology, resources, supervision, review and editing

HJF: conceptualization, funding acquisition, methodology, resources, supervision, review and editing

AL: conceptualization, funding acquisition, methodology, resources, supervision, review and editing

MK: conceptualization, funding acquisition, methodology, resources, supervision, review and editing

HT: conceptualization, funding acquisition, methodology, resources, supervision, review and editing

KP: conceptualization, formal analysis, investigation, methodology, project administration, supervision, validation, visualization, writing – original draft, and writing – review and editing

CJ: conceptualization, data curation, formal analysis, funding acquisition, investigation, methodology, project administration, resources, supervision, validation, visualization, writing – review and editing

## References

[1] Delmas, C. et al. Baseline characteristics, management, and predictors of early mortality in cardiogenic shock: insights from the frenshock registry. ESC Heart Failure 9, 408–419 (2022).

[2] Thiele, H., Ohman, E. M., de Waha-Thiele, S., Zeymer, U. & Desch, S. Management of cardiogenic shock complicating myocardial infarction: an update 2019. European Heart Journal 40, 2671–2683 (2019).

[3] Jung, R. G., et al. Prognostic factors associated with mortality in cardiogenic shock—a systematic review and meta-analysis. NEJM evidence 3, EVIDoa2300323 (2024).

[4] Zweck, E. et al. Clinical course of patients in cardiogenic shock stratified by phenotype. Heart Failure 11, 1304–1315 (2023).

[5] Naidu, S. S. et al. Scai shock stage classification expert consensus update: A review and incorporation of validation studies: This statement was endorsed by the american college of cardiology (acc), american college of emergency physicians (acep), american heart association (aha), european society of cardiology (esc) association for acute cardiovascular care (acvc), international society for heart and lung transplantation (ishlt), society of critical care medicine (sccm), and society of thoracic surgeons (sts) in december 2021. Journal of the American College of Cardiology 79, 933–946 (2022).

[6] Pöss, J. et al. Risk stratification for patients in cardiogenic shock after acute myocardial infarction. Journal of the American College of Cardiology 69, 1913–1920 (2017).

[7] Granholm, A., Møller, M. H., Krag, M., Perner, A. & Hjortrup, P. B. Predictive performance of the simplified acute physiology score (saps) ii and the initial sequential organ failure assessment (sofa) score in acutely ill intensive care patients: post-hoc analyses of the sup-icu inception cohort study. PLoS One 11, e0168948 (2016).

[8] Sleeper, L. A. et al. A severity scoring system for risk assessment of patients with cardiogenic shock: a report from the shock trial and registry. American heart journal 160, 443–450 (2010).

[9] Harjola, V.-P. et al. Clinical picture and risk prediction of short-term mortality in cardiogenic shock. European journal of heart failure 17, 501–509 (2015).

[10] Yamga, E. et al. Optimized risk score to predict mortality in patients with cardiogenic shock in the cardiac intensive care unit. Journal of the American Heart Association 12, e029232 (2023).

[11] Ceglarek, U. et al. The novel cystatin c, lactate, interleukin-6, and n-terminal pro-b-type natriuretic peptide (clip)-based mortality risk score in cardiogenic shock after acute myocardial infarction. European Heart Journal 42, 2344–2352 (2021).

[12] Stephens, A. F. et al. Ecmo pal: using deep neural networks for survival prediction in venoarterial extracorporeal membrane oxygenation. Intensive Care Medicine 49, 1090–1099 (2023).

[13] Moor, M., et al. Predicting sepsis using deep learning across international sites: a retrospective development and validation study. EClinicalMedicine 62 (2023).

[14] Zou, H. et al. Diagnosis of neurosyphilis in hiv-negative patients with syphilis: development, validation, and clinical utility of a suite of machine learning models. EClinicalMedicine 62 (2023).

[15] Bai, Z. et al. Development of a machine learning model to predict the risk of late cardiogenic shock in patients with st-segment elevation myocardial infarction. Annals of Translational Medicine 9 (2021).

[16] Chang, Y. et al. Early prediction of cardiogenic shock using machine learning. Frontiers in Cardiovascular Medicine 9, 862424 (2022).

[17] Jentzer, J. C. et al. Machine learning approaches for phenotyping in cardiogenic shock and critical illness: part 2 of 2. JACC: Advances 1, 100126 (2022).

[18] Oliveira, M., Seringa, J., Pinto, F. J., Henriques, R. & Magalhães, T. Machine learning prediction of mortality in acute myocardial infarction. BMC Medical Informatics and Decision Making 23, 1–16 (2023).

[19] Van Calster, B. et al. Calibration: the achilles heel of predictive analytics. BMC medicine 17, 1–7 (2019).

[20] Thiele, H. et al. Multivessel versus culprit lesion only percutaneous revascularization plus potential staged revascularization in patients with acute myocardial infarction complicated by cardiogenic shock: design and rationale of culprit-shock trial. American heart journal 172, 160–169 (2016).

[21] Shadbahr, T. et al. The impact of imputation quality on machine learning classifiers for datasets with missing values. Communications Medicine 3, 139 (2023).

[22] Raghunathan, T. E., Lepkowski, J. M., Van Hoewyk, J., Solenberger, P. et al. A multivariate technique for multiply imputing missing values using a sequence of regression models. Survey methodology 27, 85–96 (2001).

[23] Azur, M. J., Stuart, E. A., Frangakis, C. & Leaf, P. J. Multiple imputation by chained equations: what is it and how does it work? International journal of methods in psychiatric research 20, 40–49 (2011).

[24] Pollard, T. et al. eicu collaborative research database (version 2.0). PhysioNet 10, C2WM1R (2019).

[25] Pollard, T. J. et al. The eicu collaborative research database, a freely available multi-center database for critical care research. Scientific data 5, 1–13 (2018).

[26] Goldberger, A. L. et al. Physiobank, physiotoolkit, and physionet: components of a new research resource for complex physiologic signals. circulation 101, e215–e220 (2000).

[27] Berg, D. D., Bohula, E. A. & Morrow, D. A. Epidemiology and causes of cardiogenic shock. Current opinion in critical care 27, 401–408 (2021).

[28] Dash, M. & Liu, H. Feature selection for classification. Intelligent data analysis 1, 131–156 (1997).

[29] Churpek, M. M. et al. Quick sepsis-related organ failure assessment, systemic inflammatory response syndrome, and early warning scores for detecting clinical deterioration in infected patients outside the intensive care unit. American journal of respiratory and critical care medicine 195, 906–911 (2017).

[30] Chen, T. & Guestrin, C. Xgboost: A scalable tree boosting system. In Proceedings of the 22nd acm sigkdd international conference on knowledge discovery and data mining, 785–794 (2016).

[31] Lundberg, S. M. & Lee, S.-I. A unified approach to interpreting model predictions. In Guyon, I. et al. (eds.) Advances in Neural Information Processing Systems 30, 4765–4774 (Curran Associates, Inc., 2017).

[32] Lundberg, S. M. et al. From local explanations to global understanding with explainable ai for trees. Nature Machine Intelligence 2, 2522–5839 (2020).

[33] Štrumbelj, E. & Kononenko, I. Explaining prediction models and individual predictions with feature contributions. Knowledge and information systems 41, 647–665 (2014).

[34] Ribeiro, M. T., Singh, S. & Guestrin, C. “why should i trust you?” explaining the predictions of any classifier. In Proceedings of the 22nd ACM SIGKDD international conference on knowledge discovery and data mining, 1135–1144 (2016).

[35] Akiba, T., Sano, S., Yanase, T., Ohta, T. & Koyama, M. Optuna: A next-generation hyperparameter optimization framework. In Proceedings of the 25th ACM SIGKDD International Conference on Knowledge Discovery and Data Mining (2019).

[36] McElfresh, D., et al. When do neural nets outperform boosted trees on tabular data? arXiv preprint arXiv:2305.02997 (2023).

[37] Tolles, J. & Meurer, W. J. Logistic regression: relating patient characteristics to outcomes. Jama 316, 533–534 (2016).

[38] Valente, G., Castellanos, A. L., Hausfeld, L., De Martino, F. & Formisano, E. Cross-validation and permutations in mvpa: Validity of permutation strategies and power of cross-validation schemes. Neuroimage 238, 118145 (2021).

[39] McKinney, W. et al. Data structures for statistical computing in python. In Proceedings of the 9th Python in Science Conference, vol. 445, 51–56 (Austin, TX, 2010).

[40] Harris, C. R. et al. Array programming with numpy. Nature 585, 357– 362 (2020).

[41] Pedregosa, F. et al. Scikit-learn: Machine learning in python. the Journal of Machine Learning research 12, 2825–2830 (2011).

[42] Lemaître, G., Nogueira, F. & Aridas, C. K. Imbalanced-learn: A python toolbox to tackle the curse of imbalanced datasets in machine learning. The Journal of Machine Learning Research 18, 559–563 (2017).

[43] Virtanen, P. et al. Scipy 1.0: fundamental algorithms for scientific computing in python. Nature methods 17, 261–272 (2020).

[44] Hunter, J. D. Matplotlib: A 2d graphics environment. Computing in Science & Engineering 9, 90–95 (2007).

[45] Waskom, M. L. seaborn: statistical data visualization. Journal of Open Source Software 6, 3021 (2021).

