## Supplementary Information for "Machine learning models for early prognosis prediction in cardiogenic shock"

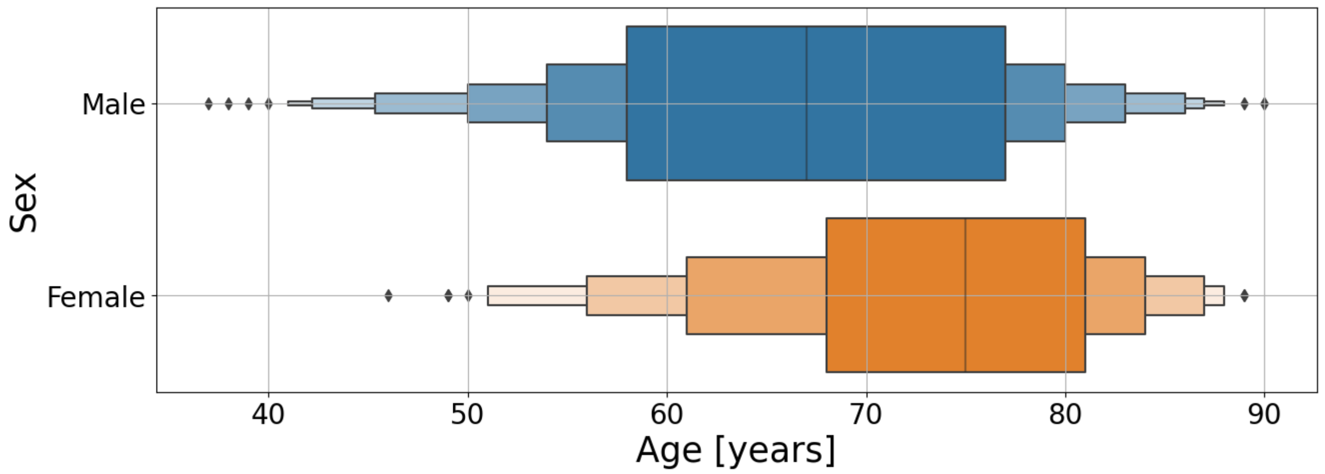


Supplementary Figure 1: Demographic distribution of patients. Age distribution was differentiated by Sex (524 male, 162 female) and 30-day outcome (361 Alive/blue, 325 Expired/orange). Each dot represents a patient in the dataset.

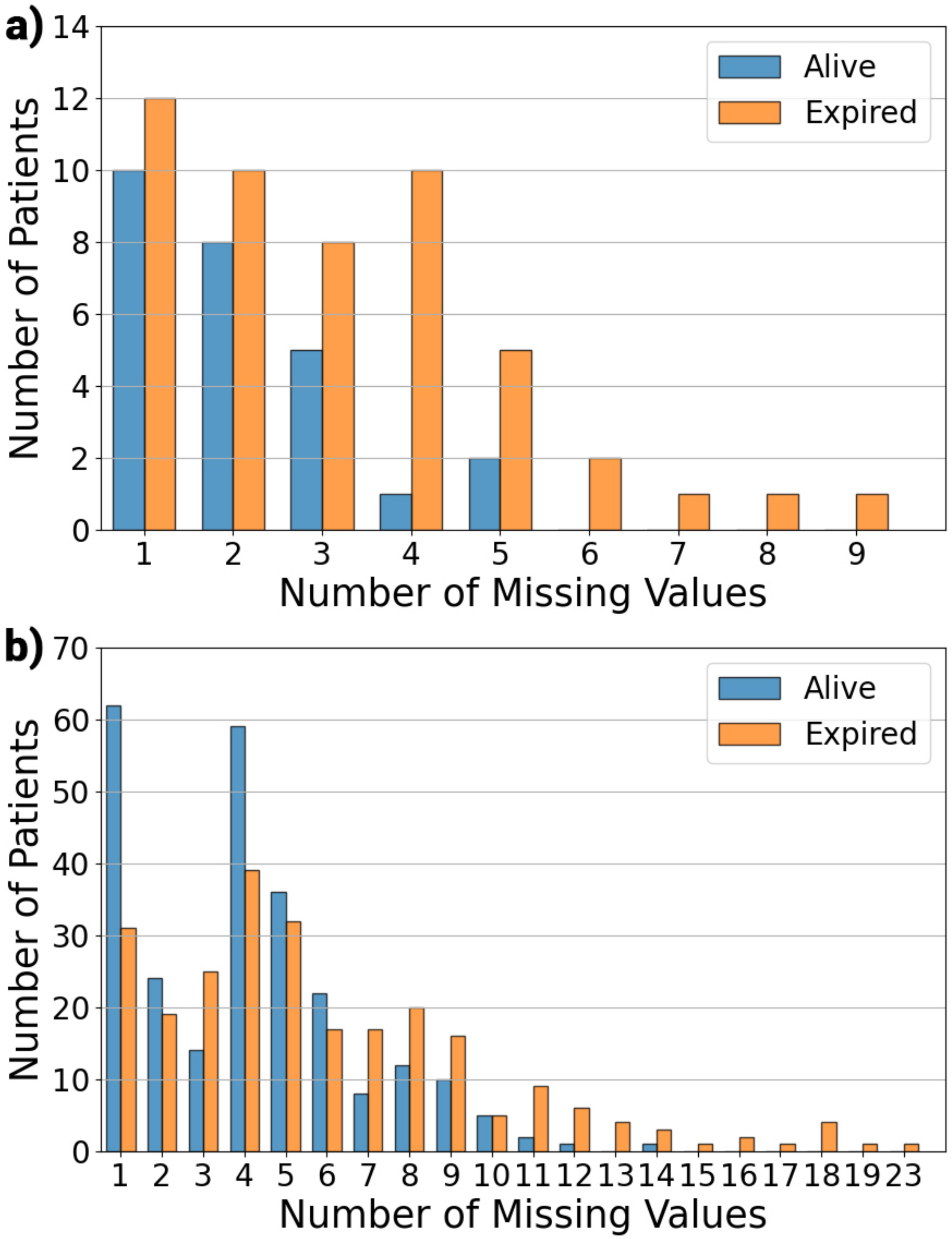


Supplementary Figure 2: Number of missing values per subject. a) Admission model. 335 Alive patients don't have missing values, while 275 Expired patients don't present any missing values in the admission features. b) Full model. 105 Alive patients don't have missing values, while 72 Expired patients don't present any missing values in the 24-hour features.

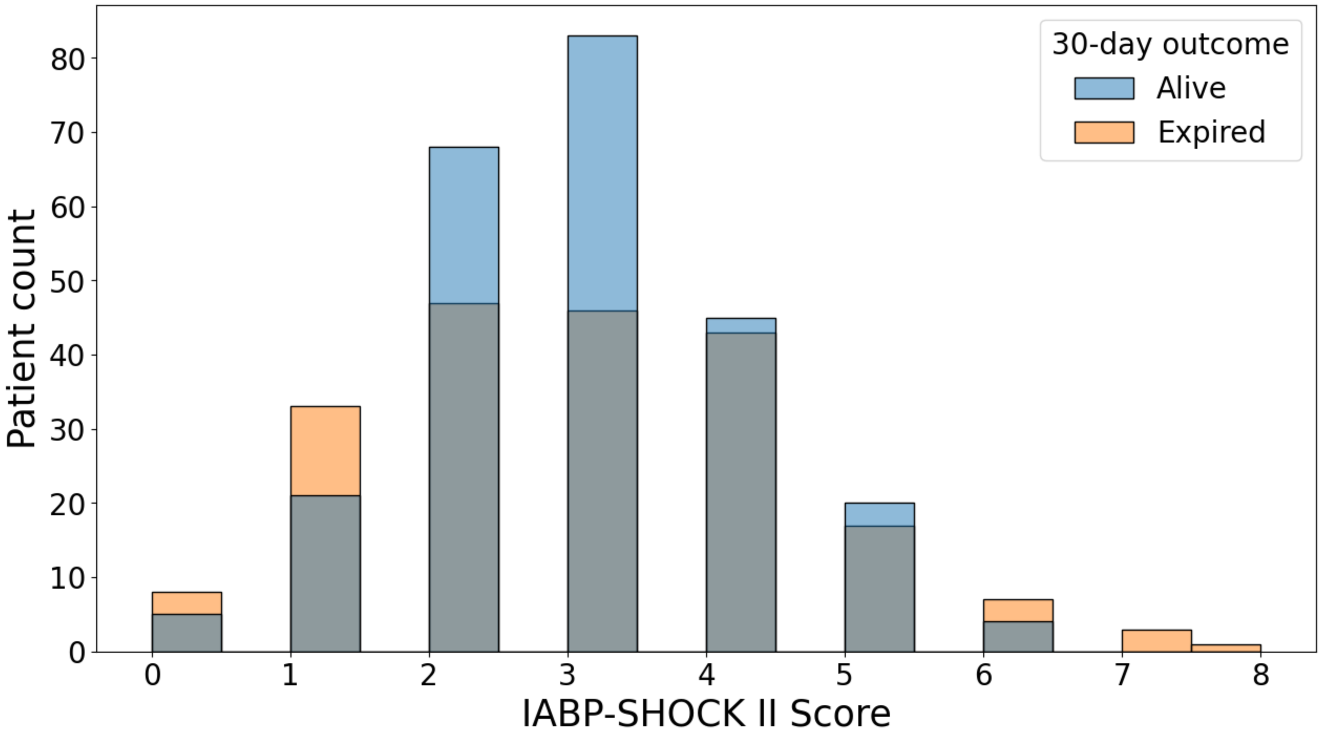


Supplementary Figure 3: IABP-SHOCK II score relationship with 30-day mortality. The risk score is not able to capture useful information about the target in the CULPRIT-SHOCK dataset.

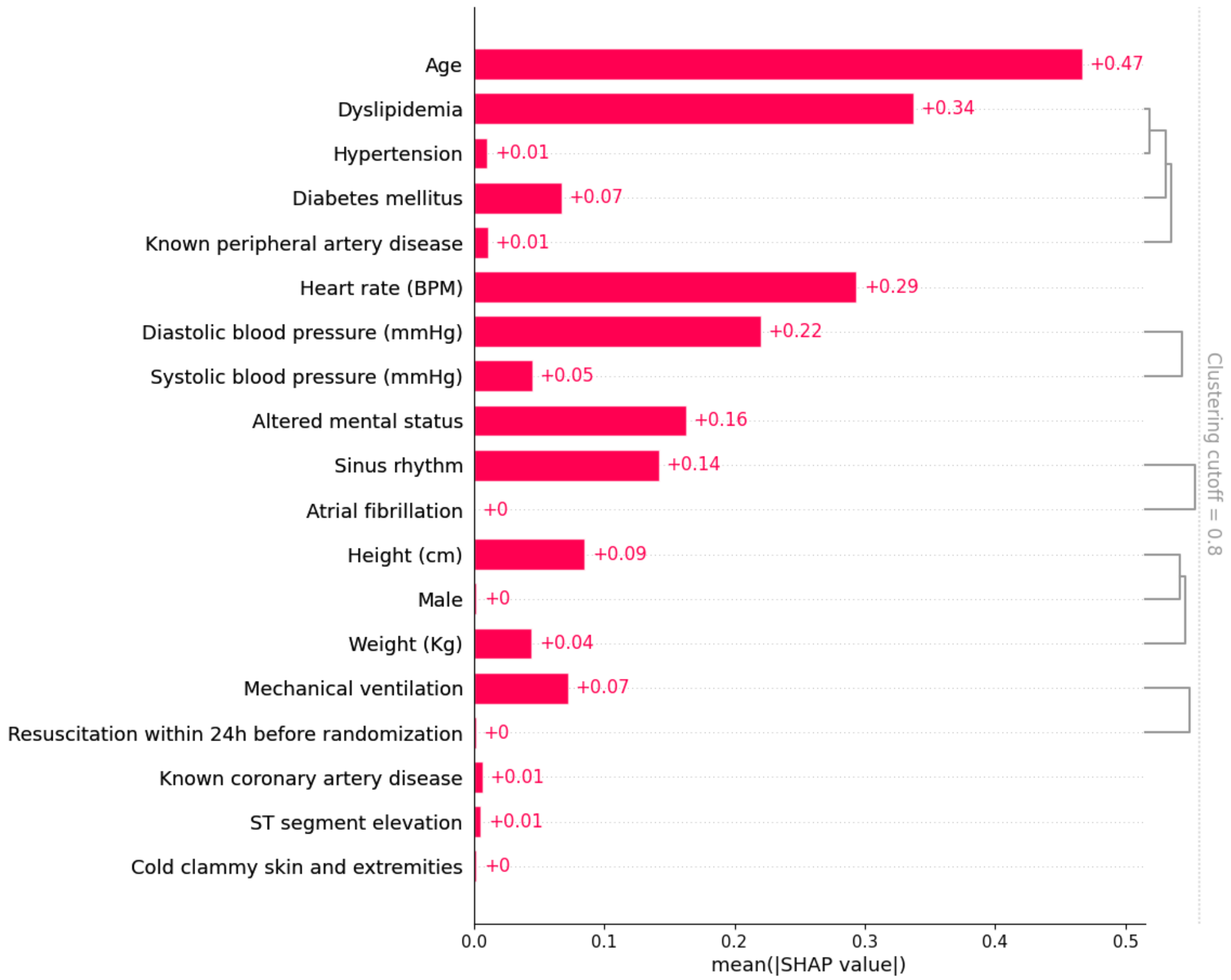


Supplementary Figure 4: Redundant feature clustering for the Admission model. The regularization aims for simpler models, making the model choose one feature if that one is highly correlated with others.

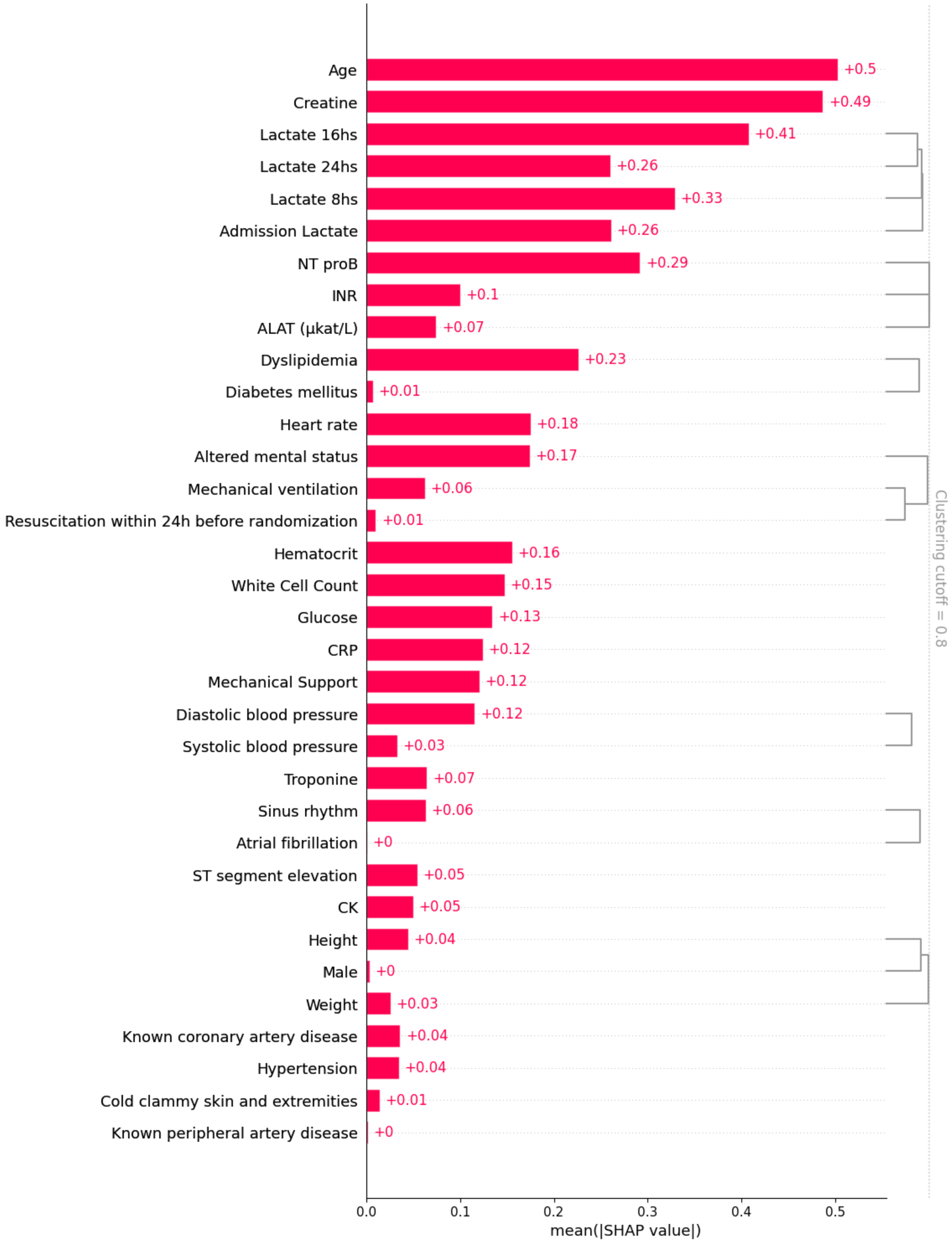


Supplementary Figure 5: Redundant feature clustering for the Full model. The regularization aims for simpler models, making the model choose one feature if that one is highly correlated with others.

Supplementary Table 1: Admission features comparison for alive and expired patients in the CULPRIT-SHOCK dataset. The values were calculated over the entire cohort.

| ID | Feature name | Mean/Count Alive | Mean/Count Expired | Missing Values Alive N-(%) | Missing Values Expired N-(%) |
|---|---|---|---|---|---|
| **Patient Information** | | | | | |
| 1 | Age [Years] | 65.55 | 71.94 | 0 (0%) | 0 (0%) |
| 2 | Sex [Male count] | 286 | 238 | 0 (0%) | 1 (1%) |
| 3 | Weight [Kg] | 82.70 | 81.72 | 5 (1%) | 13 (4%) |
| 4 | Height [cm] | 174.64 | 171.82 | 10 (2%) | 18 (5%) |
| **Comorbidities** | | | | | |
| 5 | Known coronary artery disease | 96 | 76 | 0 (0%) | 0 (0%) |
| 6 | Known peripheral artery disease | 41 | 39 | 0 (0%) | 8 (2%) |
| 7 | Arterial Hypertension | 219 | 187 | 0 (0%) | 12 (3%) |
| 8 | Dyslipidemia | 147 | 81 | 0 (0%) | 15 (4%) |
| 9 | Diabetes mellitus | 104 | 114 | 1 (1%) | 13 (4%) |
| **Electrocardiogram** | | | | | |
| 10 | Sinus rhythm | 290 | 214 | 0 (0%) | 0 (0%) |
| 11 | Atrial fibrillation | 38 | 52 | 0 (0%) | 0 (0%) |
| 12 | ST-segment elevation | 230 | 185 | 8 (2%) | 13 (4%) |
| **Clinical shock characteristics** | | | | | |
| 13 | Heart rate [bpm] | 85.30 | 91.64 | 6 (2%) | 7 (2%) |
| 14 | Systolic blood pressure [mmHg] | 101.19 | 95.14 | 6 (2%) | 9 (3%) |
| 15 | Diastolic blood pressure [mmHg] | 62.03 | 56.03 | 7 (2%) | 9 (3%) |
| 16 | Altered mental status | 223 | 238 | 1 (1%) | 3 (1%) |
| 17 | Cold, clammy skin and extremities | 237 | 232 | 7 (2%) | 6 (2%) |
| 18 | Mechanical ventilation | 206 | 202 | 0 (0%) | 4 (1%) |
| 19 | Resuscitation within 24 hours | 192 | 174 | 0 (0%) | 3 (1%) |

Supplementary Table 2: Admission features comparison for alive and expired patients in eICU dataset. The values were calculated over the entire cohort.

| ID | Feature name | Mean/Count Alive | Mean/Count Expired | Missing Values Alive N-(%) | Missing Values Expired N-(%) |
|---|---|---|---|---|---|
| **Patient Information** | | | | | |
| 1 | Age [Years] | 61.84 | 68.78 | 0 (0%) | 0 (0%) |
| 2 | Sex [Male count] | 95 | 50 | 0 (0%) | 0 (0%) |
| 3 | Weight [Kg] | 85.79 | 84.83 | 2 (1%) | 3 (4%) |
| 4 | Height [cm] | 171.16 | 169.34 | 0 (0%) | 5 (5%) |
| **Comorbidities** | | | | | |
| 5 | Known coronary artery disease | 40 | 24 | 0 (0%) | 0 (0%) |
| 6 | Known peripheral artery disease | 7 | 4 | 0 (0%) | 0 (0%) |
| 7 | Arterial Hypertension | 80 | 64 | 0 (0%) | 0 (0%) |
| 8 | Dyslipidemia | 13 | 4 | 0 (0%) | 0 (0%) |
| 9 | Diabetes mellitus | 23 | 14 | 0 (0%) | 0 (0%) |
| **Electrocardiogram** | | | | | |
| 10 | Sinus rhythm | 100 | 76 | 0 (0%) | 0 (0%) |
| 11 | Atrial fibrillation | 25 | 19 | 0 (0%) | 0 (0%) |
| 12 | ST-segment elevation | 14 | 5 | 0 (0%) | 0 (0%) |
| **Clinical shock characteristics** | | | | | |
| 13 | Heart rate [bpm] | 90.66 | 91.64 | 72 (48%) | 16 (16%) |
| 14 | Systolic blood pressure [mmHg] | 116.45 | 95.14 | 139 (93%) | 77 (80%) |
| 15 | Diastolic blood pressure [mmHg] | 62.65 | 56.03 | 139 (93%) | 78 (81%) |
| 16 | Altered mental status | 223 | 238 | 0 (0%) | 0 (0%) |
| 17 | Cold, clammy skin and extremities | 237 | 232 | 0 (0%) | 0 (0%) |
| 18 | Mechanical ventilation | 206 | 202 | 0 (0%) | 0 (0%) |
| 19 | Resuscitation within 24 hours | 192 | 174 | 0 (0%) | 0 (0%) |

Supplementary Table 3: 24 hours features comparison for alive and expired patients in CULPRIT-SHOCK dataset. The values were calculated over the entire cohort.

| ID | Feature name [unit] | Mean/Count Alive | Mean/Count Expired | Missing Values Alive N-(%) | Missing Values Expired N-(%) |
|---|---|---|---|---|---|
| **Organ perfusion** | | | | | |
| 21 | Serum lactate - Admission [mmol/L] | 3.74 | 4.68 | 47 (13%) | 52 (16%) |
| 22 | Serum lactate 8hs [mmol/L] | 2.74 | 5.02 | 55 (15%) | 114 (35%) |
| 23 | Serum lactate 16hs [mmol/L] | 1.98 | 6.14 | 66 (18%) | 143 (44%) |
| 24 | Serum lactate 24hs [mmol/L] | 1.58 | 5.08 | 80 (22%) | 161 (49%) |
| **Laboratory assessment** | | | | | |
| 25 | Nt-pro BNP [pg/ml] | 2831 | 7412 | 131 (36%) | 130 (40%) |
| 26 | CK [mmol/L] | 10200 | 17900 | 31 (8%) | 58 (17%) |
| 27 | Troponine [pg/ml] | 1542 | 5097 | 131 (63%) | 130 (40%) |
| 28 | Creatine [µmol/L] | 146.44 | 155.46 | 6 (2%) | 32 (10%) |
| 29 | White blood cell count [$10^9/L$] | 14.67 | 16.86 | 10 (2%) | 30 (9%) |
| 30 | Hematocrit [%] | 40.16 | 38.24 | 15 (4%) | 39 (12%) |
| 31 | CRP [mg/dL] | 191.44 | 260.09 | 57 (15%) | 76 (23%) |
| 32 | INR | 1.68 | 2.45 | 139 (38%) | 140 (43%) |
| 33 | Glucose [mg/dL] | 239.71 | 392.13 | 86 (23%) | 86 (26%) |
| 34 | ALAT [µkat/L] | 2.75 | 3.70 | 131 (36%) | 130 (40%) |
| **Treatment modalities** | | | | | |
| 35 | Mechanical support | 76 | 90 | 0 (0%) | 0(0%) |

Supplementary Table 4: 24 hours features comparison for alive and expired patients in eICU dataset. The values were calculated over the entire cohort.

| ID | Feature name | Mean/Count Alive | Mean/Count Expired | Missing Values Alive N-(%) | Missing Values Expired N-(%) |
|---|---|---|---|---|---|
| 21 | Serum lactate - Admission [mmol/L] | 3.89 | 7.59 | 127 (85%) | 66 (68%) |
| 22 | Serum lactate 8hs [mmol/L] | 3.90 | 6.45 | 104 (69%) | 48 (50%) |
| 23 | Serum lactate 16hs [mmol/L] | 2.43 | 5.93 | 124 (83%) | 62 (64%) |
| 24 | Serum lactate 24hs [mmol/L] | 2.49 | 6.98 | 133 (89%) | 74 (77%) |
| **Laboratory assessment** | | | | | |
| 25 | Nt-pro BNP [pg/ml] | 3174 | 4501 | 111 (74%) | 66 (68%) |
| 26 | CK [mmol/L] | 4974 | 9807 | 98 (65%) | 52 (54%) |
| 27 | Troponine [pg/ml] | 2938 | 2194 | 145 (97%) | 92 (95%) |
| 28 | Creatine [µmol/L] | 163.37 | 192.84 | 3 (2%) | (0%) |
| 29 | White blood cell count [$10^9/L$] | 13.49 | 16.90 | 9 (6%) | 4 (4%) |
| 30 | Hematocrit [%] | 35.96 | 35.05 | 9 (6%) | 3 (3%) |
| 31 | CRP [mg/dL] | 32.40 | 21.41 | 148 (99%) | 94 (97%) |
| 32 | INR | 1.62 | 1.87 | 48 (32%) | 19 (19%) |
| 33 | Glucose [mg/dL] | 161.85 | 217.08 | 3 (2%) | 0 (0%) |
| 34 | ALAT [µkat/L] | 5.11 | 8.67 | 44 (29%) | 18 (18%) |
| **Treatment modalities** | | | | | |
| 35 | Mechanical support | 31 | 17 | 118 (79%) | 78 (82%) |

Supplementary Table 5: SAPS II Scores features composition

| ID | Feature Name |
|---|---|
| 1 | Age, years |
| 2 | Heart rate |
| 3 | Systolic BP, mmHg |
| 4 | Temperature $\geq$ 39ºC (102.2ºF) |
| 5 | Glasgow Coma Scale GCS |
| 6 | $PaO_2$ / $FiO_2$, if on mechanical ventilation or Continuous positive airway pressure |
| 7 | BUN, mg/dL or serum urea, mmol/L |
| 8 | Urine output, mL/day |
| 9 | Sodium, mEq/L or mmol/L |
| 10 | Potassium, mEq/L |
| 11 | Bicarbonate, mEq/L |
| 12 | Bilirubin |
| 13 | WBC, x $10^3/mm^3$ |
| 14 | Chronic disease |
| 15 | Type of admission |

Supplementary Table 6: IABP-SHOCK II Scores Features Composition

| ID | Feature Name |
|---|---|
| 1 | Age >73 y |
| 2 | Prior stroke |
| 3 | Admission glucose >191 mg/dl |
| 4 | Creatinine >1.5 mg/dl |
| 5 | Lactate >5 mmol/l |
| 6 | Post-PCI TIMI flow grade <3 |

Supplementary Table 7: Features in CLIP score

| Feature Name | Nº of Patients with Information | Data Type | Unit |
|---|---|---|---|
| Cystatin C | 384 | Float | mg/l |
| Lactate | 374 | Float | mmol/L |
| Interleukin-6 | 384 | Float | pg/ml |
| NT pro B-type natriuretic peptide | 384 | Float | pg/ml |

Supplementary Table 8: CardShock risk score features composition. ACS, acute coronary syndrome; CABG, coronary artery bypass grafting; MI, myocardial infarction; eGFR$_{CKD-EPI}$, estimated glomerular filtration rate by the Chronic Kidney Disease Epidemiology Collaboration formula.

| ID | Feature Name | CardShock risk score |
|---|---|---|
| 1 | Age $> 75$ | 1 |
| 2 | Confusion at presentation | 1 |
| 3 | Previous MI or CABG | 1 |
| 4 | ACS aetiology | 1 |
| 5 | LVEF $< 40\%$ | 1 |
| 6 | Blood lactate | |
| | $<$ 2 mmol/L | 0 |
| | 2-4 mmol/L | 1 |
| | $>$ 4mmol/L | 2 |
| 7 | eGFR$_{CKD-EPI}$. | |
| | $>$ 60 mL/min/1.73m$^2$ | 0 |
| | 30-60 mL/min/1.73m$^2$ | 1 |
| | <3 0 mL/min/1.73m$^2$ | 2 |

Supplementary Table 9: BOS,MA$_2$ Scores Features Composition. A patient receives 1 point for meeting each of the criteria as specified in the risk score. BUN indicates blood urea nitrogen.

| ID | Feature Name | Score points |
|---|---|---|
| 1 | Maximum BUN $\geq$ 25 mg/dL | 1 |
| 2 | Minimum Oxygen saturation $<$ 88% | 1 |
| 3 | Minimum Systolic blood pressure $<$ 80 mmHg | 1 |
| 4 | Mechanical ventilation | 1 |
| 5 | Age $\geq$ 60 years | 1 |
| 6 | Maximum anion gap $\geq$ 14 mmol/L | 1 |

| Score | 0 | 1 | 2 | 3 | 4 | 5 | 6 |
|---|---|---|---|---|---|---|---|
| Risk | 0.5% | 1.4% | 3.9% | 10.0% | 23.5% | 46.0% | 70.2% |

Supplementary Table 10: Hyperparameters ranges.

| **Hyperparameter** | **Used range [min, max]** |
|---|---|
| max depth | [1, 5] |
| alpha | [1e-8, 10] |
| lambda | [1e-8, 10] |
| eta | [0.1, 1] |

Supplementary Table 11: Metrics description. All the metrics used and reported are present in this table. The formula along with the function used to compute each metric is presented. Note that the precision score is only used in the F1 score but is not reported in the results.

| **Metric** | **Formula** | **Implementation function** |
|---|---|---|
| Specificity | $\frac{TruePositive}{TrueNegative+FalsePositive}$ | imblearn.metrics.specificity_score |
| Sensitivity / Recall | $\frac{TruePositive}{TruePositive+FalseNegatives}$ | imblearn.metrics.sensitivity_score |
| Balanced Accuracy (bACC) | $\frac{Specificity+Sensitivity}{2}$ | sklearn.metrics.balanced_accuracy_score |
| Precision | $\frac{TruePositive}{TruePositive+FalsePositive}$ | sklearn.metrics.precision_score |
| F1 | $\frac{2*Precision*Sensitivity}{Precision+Sensitivity}$ | sklearn.metrics.f1_score |
| AUC | - | sklearn.metrics.roc_auc_score |
| Confidence intervals | - | stats.t.interval |

| Model | bACC (%) | AUC | F1 | Specificity | Sensitivity |
|---|---|---|---|---|---|
| Admission model (Test) | 64.6 [53.1/76.1] | 0.715 [0.598/0.832] | 0.618 [0.483/0.753] | 0.687 [0.53/0.844] | 0.605 [0.432/0.778] |
| Admission model (Train) | 74.4 [67.3/81.5] | 0.831 [0.76/0.902] | 0.725 [0.648/0.802] | 0.782 [0.719/0.845] | 0.707 [0.62/0.794] |
| Train Test difference | 9.8 | 0.116 | 0.107 | 0.95 | 0.102 |
| Full model (Test) | 70.7 [59.2/0.822] | 0.799 [0.69/0.908] | 0.633 [0.472/0.794] | 0.817 [0.704/0.93] | 0.597 [0.395/0.799] |
| Full model (Train) | 93.7 [80.8/1.066] | 0.976 [0.914/1.038] | 0.925 [0.770/1.08] | 0.97 [0.903/1.037] | 0.905 [0.715/1.095] |
| Train Test difference | 23 | 0.177 | 0.292 | 0.153 | 0.308 |

Supplementary Table 12: **Averaged results train and test from the 10 folds x 10 repetitions for the different models.** The mean and 95% confidence intervals (CI) are presented for each metric and each model. Performance on the CULPRIT-SHOCK dataset.
